# Community-Based Phenotypic Study of Safety, Tolerability, Reactogenicity and Immunogenicity of Emergency-Use-Authorized Vaccines Against COVID-19 and Viral Shedding Potential of Post-Vaccination Infections: Protocol for an Ambispective study

**DOI:** 10.1101/2021.06.28.21256779

**Authors:** Irene S. Gabashvili

**Affiliations:** Aurametrix

## Abstract

The outbreak of severe acute respiratory syndrome coronavirus 2 (SARS-CoV-2) led to a global pandemic that disrupted and impacted lives in unprecedented ways. Within less than a year after the beginning of the COVID-19 pandemic, vaccines developed by several research teams were emergency-use authorized and made their way to distribution sites across the US and other countries. COVID-19 vaccines were tested in clinical trials with thousands of participants before authorization, and were administered to over a billion people across the globe in the following 6 months. Post-authorization safety monitoring was performed using pre-existing systems (such as the World Health Organization’s platform VigiBase or US Vaccine Adverse Event Reporting System, VAERS) and newly developed post-vaccination health checkers (such as V-safe in the US). Vaccinated individuals were also posting their experiences on multiple social media groups created on Facebook, Reddit, Telegram and other platforms, but the groups were often removed as “proliferating false claims”. These forms of reporting are susceptible to biases and misclassifications and do not reach all vaccinated individuals, raising questions about risks of exacerbating health inequalities as well as security and privacy vulnerabilities.

The objective of this paper is to present the protocol for a community-based participatory research approach enabling long-term monitoring of health effects, strengthening community participation via transparent messaging and support, and addressing challenges of transitioning to a new normal.

## 2 INTRODUCTION: BACKGROUND INFORMATION AND SCIENTIFIC RATIONALE

### 2.1 Background Information

Vaccine responses vary greatly from individual to individual [1,2]. Factors that influence humoral and cellular vaccine responses include intrinsic variables such as age, sex, genetics, body fat, and comorbidities; perinatal factors such as gestational age and birth weight, extrinsic factors (such as preexisting immunity, microbiota, infections, and antibiotics), environmental factors, including nutrition, stress and timing of the vaccination, and behavioral factors such as exercise, and sleep. These variables might play differently for different vaccines and administration factors (needle length, injection technique, injecting too high or too deep in the shoulder). An understanding of the complex interplay among these factors is very important and would help to design better vaccines and vaccination strategies, reducing the risk of serious side effects.

### 2.2 Rationale

Community-based decentralized clinical trials are still a new concept. Our previous virtual solutions [3-6] laid the foundation for such studies and showed their potential and importance for the future of healthcare.

As value-based care keeps developing, community-based health is becoming one of key strategies to boost the health of a population. The involvement of community first responders (CFRs), volunteer members of the community trained to deal with medical emergencies, has expanded rapidly in recent years. Community health workers are experts in addressing the social determinants of health (SDOH) that affect specific populations (that may include individuals who do not trust the healthcare organization or do not visit the healthcare organization). Community-based approach played critical role in the COVID-19 response and can continue to be instrumental in post COVID world.

This ambispective study seeks to engage community health researchers to measure the progress and effectiveness of vaccinations and adverse events following immunization (AEFI), while helping communities safely return to pre-pandemic lives. The study includes a retrospective review to identify COVID-19 infection in 2020 and includes data from past events.

### 2.3 Potential Risks and Benefits

#### 2.3.1 Potential Risks

There are risks, discomforts, and inconveniences associated with any research study. Some survey questions may make certain participants uncomfortable. To mitigate the risk, investigators will skip questions they feel may be too sensitive. In digital surveys all questions will be optional.

Participants’ data, survey responses, and/or personally identifying information may be compromised in the event of a security breach or failure to follow protocol. To help mitigate cybersecurity breach, sensitive data will not be hosted online and investigators’ personal computers will be equipped with up-to-date antimalware software. In the event of a breach, if participants’ data are associated with their identity, they may be made public. However, since this study is decentralized, each investigator will have data for only a subset of participants and will have no access to the rest of the data.

If investigators decide to publish results from this study, de-identified participant information may be included within pooled summaries that are made public. Identification of individual-level data from those summaries would be extremely difficult, but it is possible that a third party that has obtained partial data from participants directly could compare their partial data to the published results and indirectly determine some of individual survey responses. As with any online service, if participants disclose their email account password to others, they may be able to access their account and their information. There may be additional risks to participation that are currently unforeseeable but the study will be monitored closely to minimize the possibility of any risks.

#### 2.3.2 Potential Benefits

Participants may learn about newly characterized adverse reactions to the vaccine administered to them or new treatment before it is available to everyone. Researchers may provide participants with more frequent community health check-ups as part of this research. Others may benefit from knowledge gained in this study that may aid in the development of more personalized vaccination and medical prophylaxis strategies.

OBJECTIVES AND OUTCOME MEASURES

### 2.4 Primary

| **Objective** | **Brief Description/Justification of Outcome Measure** | **Outcome Measured by** | **Time Frame** |
| --- | --- | --- | --- |
| To assess adverse reactions/events in the first hours or days following administration of the vaccine shots. | This outcome measure was chosen to assess safety and tolerability of the vaccine in the communities. | Percentage of occurrence, types, duration and severity of adverse events occurring within hours or days post-doses 1 and 2 | 10 days post doses 1 and 2 (or single dose for Janssen/Johnson & Johnson vaccine) |

Our data will be compared to local and systemic reactions (reactogenicity) of the COVID-19 vaccines reported by participants of clinical trials [7-14] and participants of v-safe or other prospective studies [15,16] 0 to 7 days after the injection. We will take into account confounding factors [17-22], previously proposed approaches [22-24] and will also review reports from social media and clinical case studies.

### 2.5 Secondary

| **Objective** | **Brief Description/Justification of Outcome Measure** | **Outcome Measured By** | **Time Frame** |
| --- | --- | --- | --- |
| To assess long-term adverse reactions/events in the 12 months following administration of the doses. | This outcome measure was chosen to assess potential long-term effects of the vaccine in different sub-cohorts. | Percentage of occurrence, types, duration and severity of adverse events occurring within months post-doses 1 and 2 | Throughout the study period, until 12 months post-final-dose |
| To assess COVID-19 infections pre- | This outcome measure was chosen to assess the duration of immunity. | Percentage of participants with COVID-19 Disease | From 14 days after completion of the 2-dose |
| and post-vaccination |  |  | regimen up to 12 months post-final-dose] |

Literature reviews of long-term and unusual side effects of COVID-19 vaccines will be performed continuously, including evaluation of dynamically evolving Adverse Events of Special Interest (AESI) and establishing background rates for these AESI [25-36]. We will also analyze breakthrough infections and efficacy of COVID-19 vaccines in different communities and populations [37-44].

## 3 STUDY DESIGN

This is a longitudinal multi-country, multi-site, decentralized, case-cohort surveillance study designed to collect both retrospective and prospective data.

The study utilizes help of multiple community investigators and assistant investigators personally acquainted with study participants in their subgroup, to continuously monitor these participants for potential health issues. No one individual in this study has access to medical histories and other personally identifiable information for all participants

The study flowchart for each investigator is as follows:

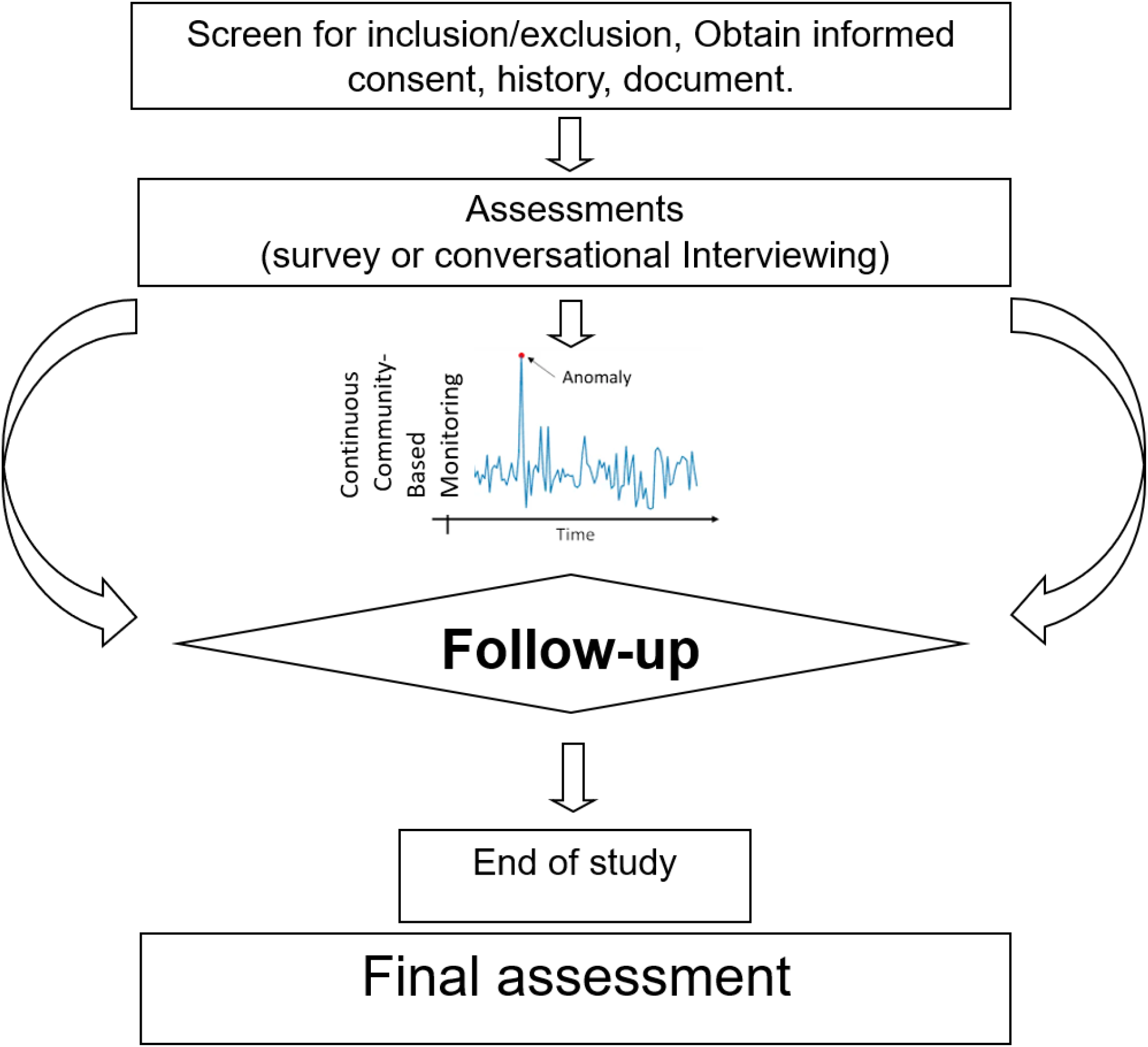

Every investigator is responsible for safely keeping identifiable participant information and shares deidentified data with the site (community) principal investigator. Site investigator has a list of all members of the community and makes sure that sub-communities are not overlapping and efforts are not duplicated. Principal investigator of the study integrates all de-identified information and regroups sub-cohorts as needed for data analysis. Investigators meet and discuss latest news and research papers, new symptoms and signs to watch for. Survey questions are modified accordingly, but every investigator personalizes them to their communities and individual participants.

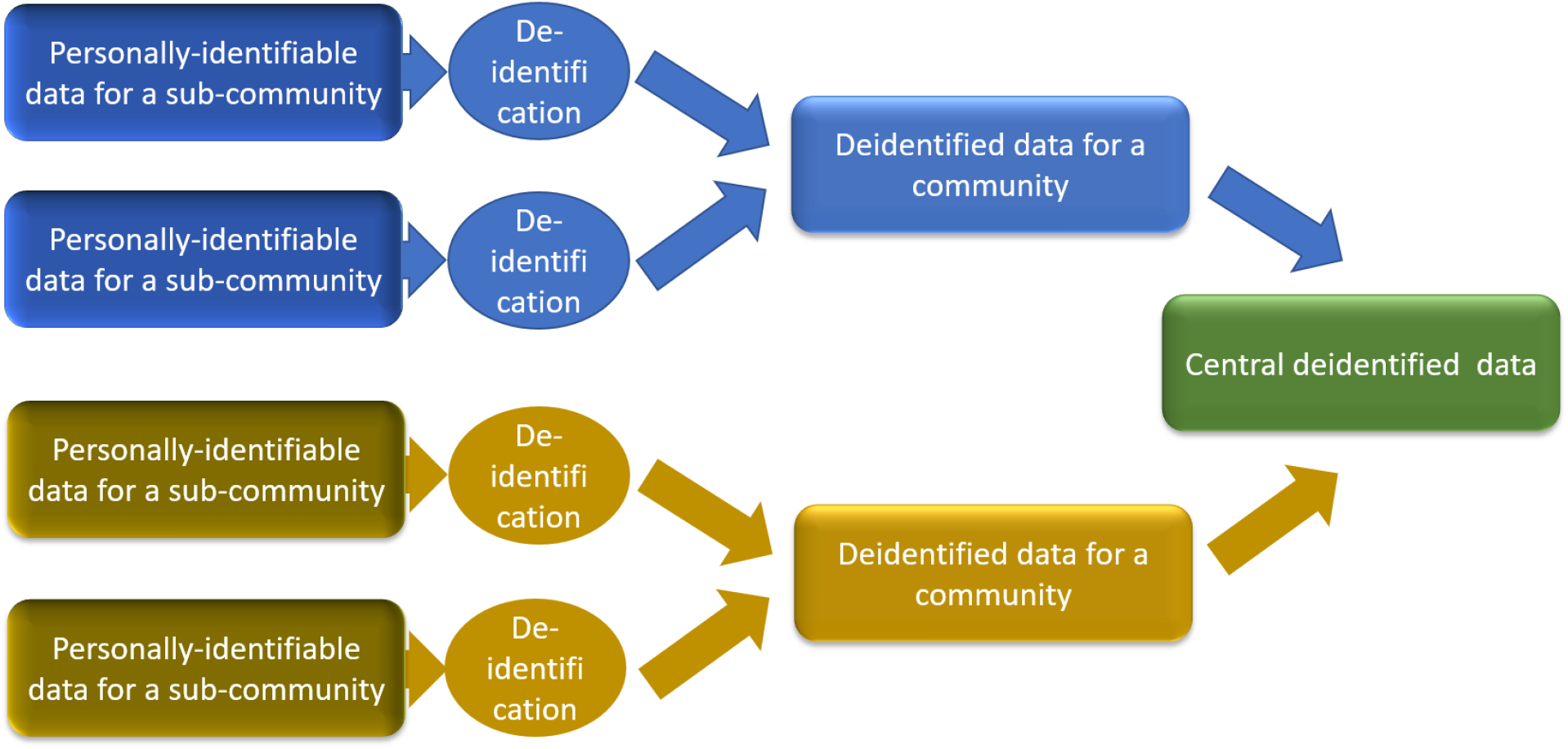

## 4 STUDY POPULATION

The population is everyone interested and eligible for emergency-use-authorized COVID-19 vaccines or interested in contributing observations to this study.

### 4.1 Participant Inclusion Criteria

- Individuals 16 or older at the time of consent
- Intention to vaccinate and/or of being available for entire study period

### 4.2 Participant Exclusion Criteria

- Any illness or condition that in the opinion of the investigator may affect the safety of the participant or the evaluation of any study endpoint.

### 4.3 Participant Withdrawal

Participants are free to withdraw from participation in the study at any time and for any reason upon request.

An investigator may withdraw a participant from the study if:

- Any medical condition, event or situation occurs such that continued participation in the study would not be in the best interest of the subject.
- The participant meets an exclusion criterion (either newly developed or not previously recognized) that precludes further study participation.

#### 4.3.1 Handling of Participant Withdrawals

Safety data will be collected if participants withdrawal occurs because of an unanticipated problem (UP) or serious adverse event (SAE).

### 4.4 Premature Termination or Suspension of Study

This study may be suspended or prematurely terminated if there is sufficient reasonable cause. The Principal Investigator is responsible for promptly notifying all parties and providing the reason(s) for the termination or suspension.

Circumstances that may warrant termination include, but are not limited to:

- Determination of unexpected, significant, or unacceptable risk to subjects.
- Insufficient adherence to protocol requirements.
- Data not sufficiently complete and/or evaluable.
- Determination of futility.

## 5 STUDY PROCEDURES/EVALUATIONS

Study data including clinical observations and/or questionnaire responses will be collected in a manner most convenient for each participant. Examples of data are:

- Medical history obtained by interview or from medical records - depending on the site.
- Medication history including a review of permitted and prohibited medications.
- Observation of participant behaviors.
- Administration of questionnaires, interviews or other instruments for subject-reported outcomes

ASSESSMENT OF SAFETY

The study procedures (interviews and questionnaires) don’t involve any known risks. Since certain subjects might experience discomfort when asked certain questions, no answers in surveys will be mandatory and the investigators will avoid or minimize the infliction of potential psychological discomfort in every individual caser.

### 5.1 Definitions of Safety Parameters

#### 5.1.1 Adverse Events

An adverse event (AE) is any untoward or unfavorable medical occurrence in a human subject, including any abnormal sign (for example, abnormal physical exam or laboratory finding), symptom, or disease, temporally associated with the subject’s participation in the research, whether or not considered related to the subject’s participation in the research.

##### 5.1.1.1 Serious Adverse Event

A serious adverse event (SAE) is one that meets one or more of the following criteria:

- Results in death
- Is life-threatening (places the subject at immediate risk of death from the event as it occurred)
- Results in inpatient hospitalization or prolongation of existing hospitalization
- Results in a persistent or significant disability or incapacity
- Results in a congenital anomaly or birth defect
- Based upon appropriate medical judgment, the event may jeopardize the subject’s health and may require medical or surgical intervention to prevent one of the other outcomes listed in this definition.

#### 5.1.2 Unanticipated Problems

The Office for Human Research Protections (OHRP) considers unanticipated problems involving risks to subjects or others to include, in general, any incident, experience, or outcome that meets all of the following criteria:

- unexpected in terms of nature, severity, or frequency given (a) the research procedures that are described in the protocol-related documents, such as the IRB-approved research protocol and informed consent document; and (b) the characteristics of the subject population being studied;
- related or possibly related to participation in the research (possibly related means there is a reasonable possibility that the incident, experience, or outcome may have been caused by the procedures involved in the research); and
- suggests that the research places subjects or others at a greater risk of harm (including physical, psychological, economic, or social harm) than was previously known or recognized.

### 5.2 Specification of Safety Parameters

Safety monitoring for this study will focus on unanticipated problems involving risks to participants, including unanticipated problems that meet the definition of a serious adverse event.

### 5.3 Reporting Procedures

#### 5.3.1 Unanticipated Problem Reporting

Incidents or events that meet the OHRP criteria for unanticipated problems require the creation and completion of an unanticipated problem report form. OHRP recommends that investigators include the following information when reporting an adverse event, or any other incident, experience, or outcome as an unanticipated problem to the IRB:

- appropriate identifying information for the research protocol, such as the title, investigator’s name, and the IRB project number;
- a detailed description of the adverse event, incident, experience, or outcome;
- an explanation of the basis for determining that the adverse event, incident, experience, or outcome represents an unanticipated problem;
- a description of any changes to the protocol or other corrective actions that have been taken or are proposed in response to the unanticipated problem.

To satisfy the requirement for prompt reporting, unanticipated problems will be reported using the following timeline:

- Unanticipated problems that are serious adverse events will be reported to the IRB within days of the investigator becoming aware of the event.
- Any other unanticipated problem will be reported to the IRB within days of the investigator becoming aware of the problem.
- All unanticipated problems should be reported to appropriate institutional officials (as required by an institution’s written reporting procedures), the supporting agency head (or designee), and OHRP within one month of the IRB’s receipt of the report of the problem from the investigator.

All unanticipated problems will be reported to the IRB.

## 6 STUDY OVERSIGHT

The principal investigator (PI) will be responsible for study oversight, including monitoring safety, ensuring that the study is conducted according to the protocol and ensuring data integrity. The PI will review the data for safety concerns and data trends at regular intervals, and will promptly submit reportable events to the IRB that arise during the conduct of the study.

## 7 STATISTICAL CONSIDERATIONS

### 7.1 Study Hypotheses

Hypothesis to be tested: The safety profile and the magnitude and durability of immune responses to the COVID-19 vaccines as well as adverse reactions depend on pre-existing health conditions, metabolism and microbiomes.

### 7.2 Sample Size Considerations

Qualitative research experts argue that there is no straightforward answer to the question of what is the best sample size for interview-based studies [45]. Some studies [46] maintain that it is sufficient to have N=20 participants in every analytically relevant participant ‘category and 50-60 people total [47]. Since a fixed sample size is not a strict require ement in this study, our sampling strategy would be to select an individual I independently of the other individuals to the sub-cohort with probability p(S i = 1). In this case the sample size is random with expectation same as n above. Recruitment will close by October 1, 2021 at which time the final study power will be determined for all outcomes under the original protocol design assumptions and the final achieved sample size. At this stage, we did not perform any formal statistical method for sample size calculation, and believe that our target sample size of at least 500 participants is satisfactory. Large sample size, as well as successful balance of known potential confounders, will provide assurance that unmeasured or unknown potential confounders will also be equally distributed across randomized treatment groups.

Wil will maintain an overall type I error rate of 0.05 or better. Target type II error rate is10%, i.e., the power of the trial is set to 90%.

### 7.3 Final Analysis Plan

In this study, primary outcome could be measured as binary (for example, presence or absence of a headache) or as ordinal data (including measurement scales). We will be using a simple 3-point scale since there are many items in the survey describing adverse effects.

Descriptive analysis will be carried out for baseline characteristics: age, gender, self-reported conditions and co-morbidities. Nominal and ordinal data will be presented by number and percentage. Continuous data will be presented depending on normal distribution with either mean and standard deviation or (in case of no normal distribution) median with range.

Inferential statistics will be performed for both primary and secondary outcomes. We will determine if data is normally distributed or not and do statistical analysis accordingly. In case of normal distribution of original or transformed (log, clr) measures we will use an unpaired t-test. In case of a non-normal distribution, we will use the non-parametric test Mann Whitney U (Wilcoxon rank sum). P values <0.05 will be considered significant.

Missing data will be described thoroughly, and will be investigated on whether it is missing completely at random (MCAR), missing at random (MAR) or not missing at random (NMAR). Unless otherwise specified, missing data will not be imputed and will be excluded from the associated analysis.

Since this is an observational study and we do not control the assignment of different types of vaccines to subjects, a difference in covariates may exist among treatment groups. Stratification (or subclassification) will be used to control for these differences in background characteristics.

SOURCE DOCUMENTS AND ACCESS TO SOURCE DATA/DOCUMENTS

Study staff will maintain appropriate medical and research records for this study, in compliance with ICH E6, Section 4.9 and regulatory and institutional requirements for the protection of confidentiality of subjects. Study staff will permit authorized representatives of regulatory agencies to examine research records for the purposes of quality assurance reviews, audits, and evaluation of the study safety, progress and data validity.

## 8 QUALITY CONTROL AND QUALITY ASSURANCE

According to the Guidelines of Good Clinical Practice (GCP) (CPMP/ICH/135/95), we are implementing and will be maintaining quality assurance and quality control systems with written Standard Operating Procedures (SOPs).Quality control will be applied to each stage of data handling.

The following steps will be taken to ensure the accuracy, consistency, completeness, and reliability of the data:

- Investigator and assistant investigator meeting(s);
- routine site monitoring;
- ongoing site communication and training;
- data management quality control checks;
- continuous data acquisition and cleaning;
- internal review of data; and
- quality control check of the final clinical study report.

In addition, the study monitor may conduct periodic audits of the study processes.

Data generated within this study will be handled according to the relevant SOPs, inspected for inconsistencies by performing validation checks.

Adverse events will be coded using the Medical Dictionary for Regulatory Activities (MedDRA). Concomitant diseases will be coded using MedDRA.

The study database, and study documentation may be subject to a Quality Assurance audit during the course of the study.

ETHICS/PROTECTION OF HUMAN SUBJECTS

### 8.1 Ethical Standard

The investigator will ensure that this study is conducted in full conformity with the principles set forth in The Belmont Report: Ethical Principles and Guidelines for the Protection of Human Subjects of Research, as drafted by the US National Commission for the Protection of Human Subjects of Biomedical and Behavioral Research (April 18, 1979) and codified in 45 CFR Part 46 and/or the ICH E6.

### 8.2 Institutional Review Board

The protocol, informed consent form(s), recruitment materials and all participant materials have been submitted to the IRB for review and approval. Approval of both the protocol and the consent form(s) have been obtained. Any amendment to the protocol will require review and approval by the IRB before the changes are implemented in the study.

### 8.3 Informed Consent Process

Informed consent is a process that is initiated prior to the individual (or a new site investigator) agreeing to participate in the study and continues throughout study participation. Extensive discussion of risks and possible benefits of study participation will be provided to participants and their families, if applicable. A consent form describing in detail the study procedures and risks will be given to the participant. Consent forms will be IRB-approved, and the participant is required to read and review the document or have the document read to him or her. The investigator or designee will explain the research study to the participant and answer any questions that may arise. The participant will sign the informed consent document prior to any study-related assessments or procedures. Participants will be given the opportunity to discuss the study with their surrogates or think about it prior to agreeing to participate. They may withdraw consent at any time throughout the course of the study. A copy of the signed informed consent document will be given to participants for their records. The rights and welfare of the participants will be protected by emphasizing to them that the quality of their clinical care will not be adversely affected if they decline to participate in this study.

The consent process will be documented in the clinical or research record.

### 8.4 Exclusion of Women, Minorities, and Specific Age Groups

Younger age group was initially excluded from the study since guidelines for vaccinations of those younger than 16 years of age were not fully developed. On May 10th, the U.S. Food and Drug Administration expanded the emergency use authorization (EUA) for the Pfizer-BioNTech COVID-19 Vaccine for the prevention of coronavirus disease 2019 (COVID-19) caused by severe acute respiratory syndrome coronavirus 2 (SARS-CoV-2) to include adolescents 12 through 15 years of age, but this study will be focusing on adult participants.

The study documentation, data, and all other information generated will be held in strict confidence. No information concerning the study or the data will be released to any unauthorized third party without prior written approval of the study monitor.

## 9 DATA HANDLING AND RECORD KEEPING

The investigators are responsible for ensuring the accuracy, completeness, legibility, and timeliness of the data reported. All source documents should be completed in a neat, legible manner to ensure accurate interpretation of data. The investigators will maintain adequate case histories of study participants, including accurate case report forms (CRFs), and source documentation.

### 9.1 Data Management Responsibilities

Data collection and accurate documentation are the responsibility of the study staff under the supervision of the Principal Investigator. All source documents and laboratory reports must be reviewed by the study team and data entry staff, who will ensure that they are accurate and complete. Unanticipated problems must be reviewed by the Principal Investigator or designee.

### 9.2 Data Capture Methods

Every investigator will use ongoing processing methods specific to their site/subgroup including paper and electronic. Cumulative data will be maintained in electronic format. Sensitive data with patient information will be kept in decentralized fashion - password-protected on the investigator’s computer or locked in the investigator’s desk.

### 9.3 Schedule and Content of Reports

Investigators will share information weekly or more frequently, if needed.

### 9.4 Study Records Retention

Study records will be maintained for one to three years.

### 9.5 Protocol Deviations

A protocol deviation is any change, divergence, or departure from the study procedures described in the IRB-approved clinical study protocol. The deviation may be on the part of the participant, the investigator, or study staff.

Consistent with the investigator obligations in the ICH E6 Guideline for Good Clinical Practice, the Principal Investigator will document in study source documents and explain any deviation from the IRB-approved protocol. The PI will report to the IRB any deviations or changes made to eliminate immediate hazards to participants and any changes that increase risk to participants and/or significantly affect the conduct of the study.

Protocol deviations will be assessed for their impact on safety, study operations, and data integrity. Appropriate corrective and preventive actions will be implemented if warranted.

## 10 PUBLICATION/DATA SHARING POLICY

This study will comply with all applicable NIH Data Sharing Policies. See https://grants.nih.gov/policy/sharing.htm for policies and resources.

## Data Availability

The data collected in this study will be made available anonymized to other academic researchers. For more details see https://aurametrix.com/Studies/NCT04832932.html

## LITERATURE REFERENCES

1. Zimmermann P, Curtis N. Factors that influence the immune response to vaccination. Clinical microbiology reviews. 2019 Mar 20;32(2).

2. Hervé C, Laupèze B, Del Giudice G, Didierlaurent AM, Da Silva FT. The how’s and what’s of vaccine reactogenicity. npj Vaccines. 2019 Sep 24;4(1):1–1.

3. Gabashvili IS. Cutaneous Bacteria in the Gut Microbiome as Biomarkers of Systemic Malodor and People Are Allergic to Me (PATM) Conditions: Insights From a Virtually Conducted Clinical Trial. JMIR Dermatology. 2020 Nov 4;3(1):e10508.

4. Gabashvili IS. Effects of diet, activities, environmental exposures and trimethylamine metabolism on alveolar breath compounds: protocol for a retrospective case-cohort observational study. medRxiv. 2021 Jan 26.

5. Gabashvili IS. Community-led research discovers links between elusive symptoms and clinical tests. bioRxiv. 2017 May 19:139014.

6. Gabashvili IS. Identifying subtypes of a stigmatized medical condition. medRxiv. 2019 Aug 29:19005223.

7. Baden LR, El Sahly HM, Essink B, Kotloff K, Frey S, Novak R, Diemert D, Spector SA, Rouphael N, Creech CB, McGettigan J. Efficacy and Safety of the mRNA-1273 SARS-CoV-2 Vaccine. New England Journal of Medicine. 2020 Dec 30. [Moderna]\

8. Polack FP, Thomas SJ, Kitchin N, Absalon J, Gurtman A, Lockhart S, Perez JL, Pérez Marc G, Moreira ED, Zerbini C, Bailey R. Safety and efficacy of the BNT162b2 mRNA Covid-19 vaccine. New England Journal of Medicine. 2020 Dec 31;383(27):2603-15. [Pfizer/BioNTech]

9. Voysey M, Clemens SA, Madhi SA, Weckx LY, Folegatti PM, Aley PK, Angus B, Baillie VL, Barnabas SL, Bhorat QE, Bibi S. Safety and efficacy of the ChAdOx1 nCoV-19 vaccine (AZD1222) against SARS-CoV-2: an interim analysis of four randomised controlled trials in Brazil, South Africa, and the UK. The Lancet. 2021 Jan 9;397(10269):99–111. [Astrazeneca]

10. Voysey, Merryn and Costa Clemens, Sue Ann and Madhi Shabir A. and Weckx Lily Yin and Folegatti Pedro M. and Aley Parvinder K. and Angus Brian John and Baillie, Vicky and Barnabas Shaun L. and Bhorat Qasim E. and Bibi, Sagida and Briner, Carmen and Cicconi, Paola and Clutterbuck, Elizabeth and Collins Andrea M. and Cutland, Clare and Darton, Thomas and Dheda, Keertan and Douglas Alexander D. and Duncan Christopher J. A. and Emary Katherine R. W. and Ewer, Katie and Flaxman, Amy and Fairlie, Lee and Faust Saul N. and Feng, Shuo and Ferreira Daniela M. and Finn, Adam and Galiza, Eva and Goodman Anna L. and Green Catherine M. and Green Christopher A. and Greenland, Melanie and Hill, Catherine and Hill Helen C. and Hirsch, Ian and Izu, Alane and Jenkin, Daniel and Kerridge, Simon and Koen, Anthonet and Kwatra, Gaurav and Lazarus, Rajeka and Libri, Vincenzo and Lillie Patrick J. and Marchevsky Natalie G. and Marshall Richard P. and Mendes Ana Verena Almeida and Milan Eveline P. and Minassian Angela M. and McGregor Alastair C. and Farooq Mujadidi Yama and Nana, Anusha and Payadachee Sherman D. and Phillips Daniel J. and Pittella, Ana and Plested, Emma and Pollock Katrina M. and Ramasamy Maheshi N. and Robinson, Hannah and Schwarzbold Alexandre V. and Smith, Andrew and Song, Rinn and Snape Matthew D. and Sprinz, Eduardo and Sutherland Rebecca K. and Thomson Emma C. and Torok, Mili and Toshner, Mark and Turner David P. J. and Vekemans, Johan and Villafana Tonya L. and White, Thomas and Williams Christopher J. and Hill Adrian V. S. and Lambe, Teresa and Gilbert Sarah C. and Pollard, Andrew and Group, Oxford COVID Vaccine Trial, Single Dose Administration, And The Influence Of The Timing Of The Booster Dose On Immunogenicity and Efficacy Of ChAdOx1 nCoV-19 (AZD1222) Vaccine. Available at SSRN: https://ssrn.com/abstract=3777268 x[Astrazeneca single-dose]

11. Logunov DY, Dolzhikova IV, Zubkova OV, Tukhvatullin AI, Shcheblyakov DV, Dzharullaeva AS, Grousova DM, Erokhova AS, Kovyrshina AV, Botikov AG, Izhaeva FM. Safety and immunogenicity of an rAd26 and rAd5 vector-based heterologous prime-boost COVID-19 vaccine in two formulations: two open, non-randomised phase 1/2 studies from Russia. The Lancet. 2020 Sep 26;396(10255):887–97. [Sputnik]

12. Zhang Y, Zeng G, Pan H, Li C, Hu Y, Chu K, Han W, Chen Z, Tang R, Yin W, Chen X. Safety, tolerability, and immunogenicity of an inactivated SARS-CoV-2 vaccine in healthy adults aged 18–59 years: a randomised, double-blind, placebo-controlled, phase 1/2 clinical trial. The Lancet infectious diseases. 2021 Feb 1;21(2):181–92. [CoronaVac, Sinovac]

13. Wu Z, Hu Y, Xu M, Chen Z, Yang W, Jiang Z, Li M, Jin H, Cui G, Chen P, Wang L. Safety, tolerability, and immunogenicity of an inactivated SARS-CoV-2 vaccine (CoronaVac) in healthy adults aged 60 years and older: a randomised, double-blind, placebo-controlled, phase 1/2 clinical trial. The Lancet Infectious Diseases. 2021 Jun 1;21(6):803–12. [BBIBP-CorV, Sinopharm]

14. McDonald I, Murray SM, Reynolds CJ, Altmann DM, Boyton RJ. Comparative systematic review and meta-analysis of reactogenicity, immunogenicity and efficacy of vaccines against SARS-CoV-2. npj Vaccines. 2021 May 13;6(1):1–4.

15. Chapin-Bardales J, Gee J, Myers T. Reactogenicity following receipt of mRNA-based COVID-19 vaccines. Jama. 2021 Jun 1;325(21):2201-2. [v-safe, Pfizer/BioNTech, Moderna]

16. Mathioudakis, A.G.; Ghrew, M.; Ustianowski, A.; Ahmad, S.; Borrow, R.; Papavasileiou, L.P.; Petrakis, D.; Bakerly, N.D. Self-Reported Real-World Safety and Reactogenicity of COVID-19 Vaccines: A Vaccine Recipient Survey. Life 2021, 11, 249. https://doi.org/10.3390/life11030249

17. Remmel A. Why is it so hard to investigate the rare side effects of COVID vaccines? Nature. 2021 Apr 1.

18. Presby D, Capodilupo E. Objective and Subjective COVID-19 Vaccine Reactogenicity by Age and Vaccine Manufacturer. medRxiv. 2021 April 30.

19. Edwards KM, Booy R. Effects of exercise on vaccine-induced immune responses. Human vaccines & immunotherapeutics. 2013 Apr 1;9(4):907–10.

20. Kimball BA, Opiekun M, Yamazaki K, Beauchamp GK. Immunization alters body odor. Physiology & behavior. 2014 Apr 10;128:80–5.

21. Gordon AR, Kimball BA, Sorjonen K, Karshikoff B, Axelsson J, Lekander M, Lundström JN, Olsson MJ. Detection of inflammation via volatile cues in human urine. Chemical senses. 2018 Nov 1;43(9):711–9.

22. Remschmidt C, Wichmann O, Harder T. Frequency and impact of confounding by indication and healthy vaccinee bias in observational studies assessing influenza vaccine effectiveness: a systematic review. BMC infectious diseases. 2015 Dec;15(1):1–5.

23. Van Tilbeurgh M, Lemdani K, Beignon AS, Chapon C, Tchitchek N, Cheraitia L, Marcos Lopez E, Pascal Q, Le Grand R, Maisonnasse P, Manet C. Predictive Markers of Immunogenicity and Efficacy for Human Vaccines. Vaccines. 2021 Jun;9(6):579.

24. Gonzalez-Dias P, Lee EK, Sorgi S, de Lima DS, Urbanski AH, Silveira EL, Nakaya HI. Methods for predicting vaccine immunogenicity and reactogenicity. Human vaccines & immunotherapeutics. 2020 Feb 1;16(2):269–76.

25. Black SB, Law B, Chen RT, Dekker CL, Sturkenboom M, Huang WT, Gurwith M, Poland G. The critical role of background rates of possible adverse events in the assessment of COVID-19 vaccine safety. Vaccine. 2021 May 6;39(19):2712–8.

26. Tobaiqy M, Elkout H, MacLure K. Analysis of Thrombotic Adverse Reactions of COVID-19 AstraZeneca Vaccine reported to EudraVigilance database. Vaccines. 2021 Apr;9(4):393.

27. Dutta S, Kaur RJ, Charan J, Bhardwaj P, Sharma P, Ambwani S, Haque M, Tandon A, Jha P, Sukhija S, Suman SV. Serious adverse events reported from the COVID-19 vaccines: A descriptive study based on WHO database. medRxiv. 2021 March 24.

28. Raw RK, Kelly C, Rees J, Wroe C, Chadwick DR. Previous COVID-19 infection but not Long-COVID is associated with increased adverse events following BNT162b2/Pfizer vaccination. medRxiv. 2021 April 22

29. Davido B, Mascitti H, Fortier-Beaulieu M, Jaffal K, de Truchis P. ‘Blue toes’ following vaccination with the BNT162b2 mRNA COVID-19 vaccine. Journal of Travel Medicine. 2021 Feb 23.

30. Jedlowski PM, Jedlowski MF. Morbilliform rash after administration of Pfizer-BioNTech COVID-19 mRNA vaccine. Dermatology online journal. 2021;27(1).

31. Waheed S, Bayas A, Hindi F, Rizvi Z, Espinosa PS. Neurological complications of COVID-19: Guillain-Barre syndrome following Pfizer COVID-19 vaccine. Cureus. 2021 Feb;13(2).

32. Lei Y, Zhang J, Schiavon CR, He M, Chen L, Shen H, Zhang Y, Yin Q, Cho Y, Andrade L, Shadel GS. SARS-CoV-2 Spike Protein Impairs Endothelial Function via Downregulation of ACE 2. Circulation Research. 2021 Apr 30;128(9):1323–6.

33. Botwin G, Li D, Figueiredo J, Cheng S, Braun J, McGovern DP, Melmed G. Adverse Events Following SARS-CoV-2 mRNA Vaccination Among Patients with Inflammatory Bowel Disease. medRxiv. 2021 March 31.

34. Ledford H. COVID vaccines and blood clots: five key questions. Nature. 2021 Apr 16;592(7855):495–6.

35. Nassar M, Chung H, Dhayaparan Y, Nyein A, Acevedo BJ, Chicos C, Zheng D, Barras M, Mohamed M, Alfishawy M, Nso N. COVID-19 vaccine induced rhabdomyolysis: Case report with literature review. Diabetes & Metabolic Syndrome. 2021 Jun 15.

36. Enghard P, Hardenberg JH, Stockmann H, Hinze C, Eckardt KU, Schmidt-Ott KM. Long-term effects of COVID-19 on kidney function. The Lancet. 2021 May 15;397(10287):1806–7.

37. Walach, H.; Klement, R.J.; Aukema, W. The Safety of COVID-19 Vaccinations—We Should Rethink the Policy. Vaccines 2021, 9, 693. https://doi.org/10.3390/vaccines9070693

38. Daniel W, Nivet M, Warner J, Podolsky DK. Early evidence of the effect of SARS-CoV-2 vaccine at one medical center. New England Journal of Medicine. 2021 Mar 23. [NT162b2 by Pfizer–BioNTech, mRNA-1273 by Moderna]

39. Thompson, M.G., 2021. Interim Estimates of Vaccine Effectiveness of BNT162b2 and mRNA-1273 COVID-19 Vaccines in Preventing SARS-CoV-2 Infection Among Health Care Personnel, First Responders, and Other Essential and Frontline Workers—Eight US Locations, December 2020–March 2021. MMWR. Morbidity and Mortality Weekly Report, 70.

40. Levine-Tiefenbrun M, Yelin I, Katz R, Herzel E, Golan Z, Schreiber L, Wolf T, Nadler V, Ben-Tov A, Kuint J, Gazit S. Initial report of decreased SARS-CoV-2 viral load after inoculation with the BNT162b2 vaccine. Nature Medicine. 2021 Mar 29:1–3.

41. Class J, Dangi T, Richner J, Penaloza-MacMaster P. A SARS CoV-2 nucleocapsid vaccine protects against distal viral dissemination. bioRxiv. 2021 Jan 1.

42. COVID C, Team VB, COVID C, Team VB, COVID C, Team VB, Birhane M, Bressler S, Chang G, Clark T, Dorough L. COVID-19 Vaccine Breakthrough Infections Reported to CDC—United States, January 1–April 30, 2021. Morbidity and Mortality Weekly Report. 2021 May 28;70(21):792.

43. Hacisuleyman E, Hale C, Saito Y, Blachere NE, Bergh M, Conlon EG, Schaefer-Babajew DJ, DaSilva J, Muecksch F, Gaebler C, Lifton R. Vaccine breakthrough infections with SARS-CoV-2 variants. New England Journal of Medicine. 2021 Jun 10;384(23):2212–8.

44. Esteban Ramirez, Rebecca P. Wilkes, Giovanna Carpi, Jack Dorman, Craig Bowen, Lisa Smith. SARS-CoV-2 Breakthrough Infections in Fully Vaccinated Individuals. medRxiv 2021.06.21.21258990; doi: https://doi.org/10.1101/2021.06.21.21258990

45. Vasileiou K, Barnett J, Thorpe S, Young T. Characterising and justifying sample size sufficiency in interview-based studies: systematic analysis of qualitative health research over a 15-year period. BMC medical research methodology. 2018 Dec;18(1):1–8.

46. Green J, Thorogood N. Qualitative methods for health research. sage; 2018 Feb 26.

47. Britten N. Qualitative research: qualitative interviews in medical research. Bmj. 1995 Jul 22;311(6999):251–3.

